# DYNAMICS OF C-REACTIVE PROTEIN IN THE EARLY POSTOPERATIVE PERIOD AS A PREDICTOR OF INFECTIOUS COMPLICATIONS AND A TOOL FOR OPTIMIZING ANTIBIOTIC THERAPY

**DOI:** 10.64898/2026.04.06.26350253

**Authors:** Irina N. Ochakovskaya, Vladimir V. Onopriev, Narine M. Dovlatbekyan, Ksenia Sh. Zhuravleva, Georgiy Yu. Zamulin, Vladimir M. Durleshter

## Abstract

**Objective:** To evaluate the diagnostic and prognostic significance of C-reactive protein (CRP) level dynamics within the first five days after surgery for the early detection of surgical site infections (SSI) and to identify independent risk factors, taking into account regional specifics of surgical management (types of surgeries, duration of procedures), as well as the local hospital microbial landscape.

**Materials and Methods:** A single-center retrospective cohort analysis of data from 127 patients who underwent surgical procedures between 2022 and 2024 was conducted. CRP levels on postoperative days 1, 3, and 5 were assessed, and delta values (ΔCRP) were calculated. Descriptive statistics, ROC analysis, and multivariate logistic regression were used to identify predictors of SSI.

**Results:** Patients with SSI lacked the physiological decrease in CRP levels by day 5. The most informative indicator was the CRP level on day 3: a threshold of >106 mg/L was associated with a high risk of SSI (AUC=0.76; sensitivity 85%, specificity 63%). Independent predictors of SSI included surgery duration (OR=1.015 per 1 min; p<0.001) and the increase in CRP between days 3 and 5 (ΔCRP3–5: OR=1.027; p=0.023). A combined model (clinical parameters + CRP) demonstrated the highest predictive ability (AUC=0.79).

**Conclusion:** Monitoring CRP dynamics, particularly on days 3 and 5, is a highly informative and accessible method for the early diagnosis of SSI. A CRP threshold of >100 mg/L on day 3 and its subsequent increase should serve as a trigger for in-depth diagnostic investigation and rationalization of antimicrobial therapy.

## Introduction

C-reactive protein (CRP) is a classic acute-phase inflammatory marker whose synthesis is predominantly stimulated by interleukin-6 (IL-6), produced by macrophages and T-cells in response to infectious agents or tissue damage, leading to a rapid increase in its plasma level within 6–12 hours after the stimulus. In the postoperative period, CRP levels characteristically rise, reaching peak values by 48–72 hours (day 3) due to surgical trauma, followed by a decline in uncomplicated cases [1, 2]. Unlike sterile inflammation, where CRP dynamics are characterized by a rapid decline, in bacterial infection, CRP levels continue to rise or remain persistently elevated after day 3, driven by pathophysiological mechanisms such as complement activation and pathogen opsonization [3–5].

The primary clinical challenge lies in the differential diagnosis between the expected CRP elevation due to surgical trauma and its pathological rise caused by infection. This requires comparison with other markers, such as procalcitonin or the neutrophil-to-lymphocyte ratio (NLR), where CRP shows advantages in accessibility and cost but lower specificity for bacterial infections [6]. Unjustified initiation or escalation of antibiotic therapy (ABT) “on day 3” due to overdiagnosis contributes to the growth of antimicrobial resistance, which is particularly relevant in the global context where, according to the World Health Organization (WHO), antibiotic resistance causes 700,000 deaths annually and may reach 10 million by 2050, underscoring the need for Antimicrobial Stewardship (AMS) strategies [7]. In this regard, determining precise diagnostic and prognostic CRP thresholds, as well as their dynamics, is a pressing task in contemporary perioperative medicine, especially in regional studies where local specifics of patient management (types of surgeries, duration of procedures), as well as the hospital microbial landscape, allow the identification of new trends, such as variations in threshold values [4, 5, 8].

### Objective

to evaluate the diagnostic significance of absolute values and dynamics of CRP levels on postoperative days 1, 3, and 5 for the early detection of SSI, and to identify independent clinical and laboratory predictors of the development of these complications.

## Materials and Methods

### Study design

A single-center retrospective cohort study was conducted at the Regional Clinical Hospital No. 2 (Krasnodar, Russia).

### Inclusion and exclusion criteria

The analysis included data from 127 patients who underwent elective and emergency surgical procedures of various profiles (general surgery, oncology, urology) between 2022 and 2024. Inclusion criteria: availability of CRP plasma level results on postoperative days 1, 3, and 5. Exclusion criteria: presence of active systemic infection in the preoperative period (Altemeier wound class IV), immunosuppressive therapy, incomplete medical records.

### Methods

SSI was diagnosed according to CDC criteria (2017) [9, 10]. For each patient, the following clinical and demographic data were recorded: age, sex, BMI, ASA score, Altemeier wound class (I–III), surgery duration (min), and duration of perioperative antibiotic prophylaxis (PAP), which included standard regimens such as cefazolin and protected aminopenicillins in accordance with ASHP guidelines [11]. Absolute CRP values (mg/L) on days 1 (CRP1), 3 (CRP3), and 5 (CRP5) were assessed, along with calculated dynamic indicators: ΔCRP1–3 (CRP3 – CRP1), ΔCRP1–5 (CRP5 – CRP1), ΔCRP3–5 (CRP5 – CRP3).

### Statistical analysis

The Mann–Whitney U test was used for comparison of quantitative variables between groups. Diagnostic accuracy of individual parameters was assessed using ROC analysis with calculation of the area under the curve (AUC), optimal threshold (Youden index), sensitivity, and specificity. Multivariate logistic regression was used to identify independent predictors of SSI. The predictive ability of clinical, laboratory, and combined models was evaluated using 5-fold stratified cross-validation with AUC calculation and calibration assessment (calibration curves, Brier score). Statistical significance was set at p<0.05. Analysis was performed using Python 3.11 (scipy, statsmodels, scikit-learn libraries). Post-hoc power analysis using the statsmodels and pwr libraries showed that with a sample size of n=127 and an effect size of Cohen’s d=0.5, the power for group comparison was 0.82, which is acceptable for detecting moderate differences; for regression with 5 predictors, power was estimated at 0.75 at alpha=0.05. Additional subgroup analysis by surgery type (elective and emergency) and confounders (comorbidities, including diabetes and smoking, as well as microbiological infection profiles) was performed to account for potential bias and regional variations.

## Results

Of 127 patients with a complete CRP dataset, SSI was diagnosed in 21 (16.5%); in 106 patients, the postoperative course was uneventful.

### CRP Dynamics

In the group without complications, a typical CRP pattern was observed with a peak on day 3 and subsequent significant decline by day 5. In the SSI group, median CRP levels were significantly higher on days 3 and 5, and no decline by day 5 was observed (Table 1, Fig. 1).

**Table 1.**
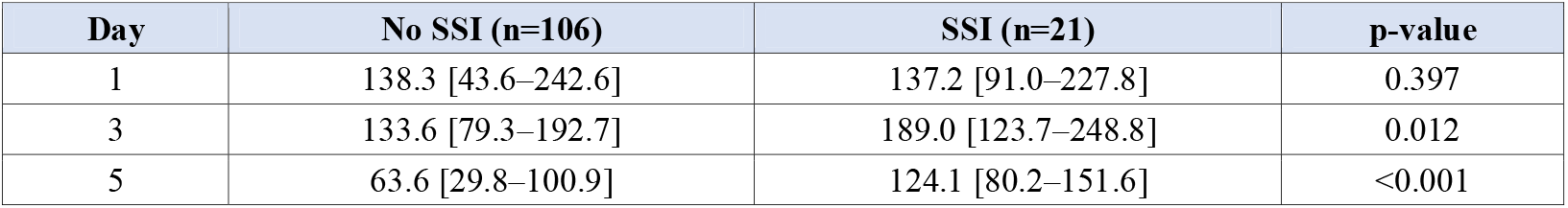
CRP levels (median [IQR]) in patients with and without SSI.

| Day | No SSI (n=106) | SSI (n=21) | p-value |
| --- | --- | --- | --- |
| 1 | 138.3 [43.6–242.6] | 137.2 [91.0–227.8] | 0.397 |
| 3 | 133.6 [79.3–192.7] | 189.0 [123.7–248.8] | 0.012 |
| 5 | 63.6 [29.8–100.9] | 124.1 [80.2–151.6] | <0.001 |

**Figure 1.** Median CRP dynamics in the postoperative period in patients with and without SSI. Note: Figure 1 (CRP dynamics plot) is from the original Russian manuscript. For final medRxiv submission, this figure should be recreated with English axis labels.

### Diagnostic Accuracy of CRP

ROC analysis demonstrated that the CRP level on day 3 had the highest predictive ability for SSI diagnosis (AUC=0.76; 95% CI: 0.67–0.85) (Fig. 2).

**Figure 2.**
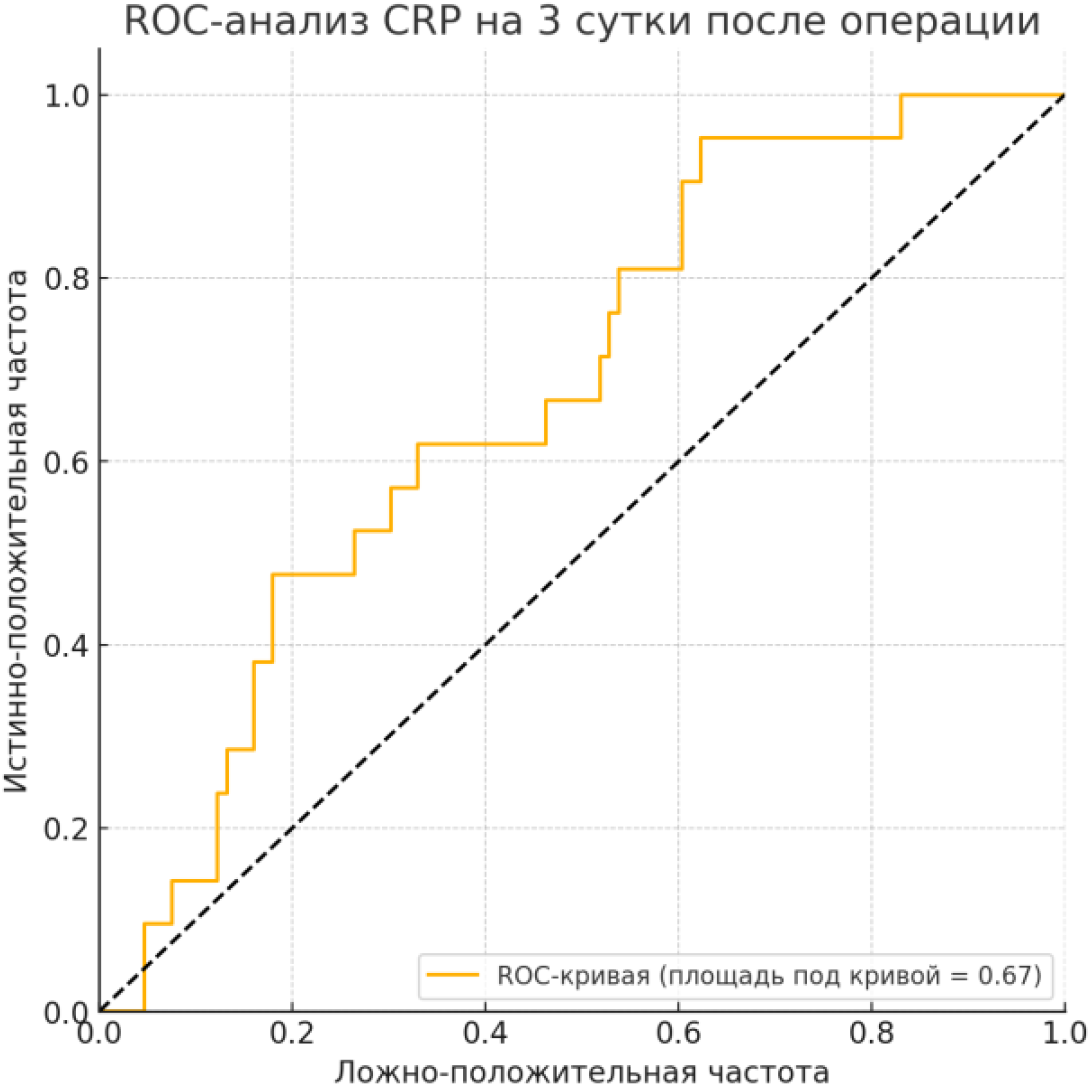
ROC curve for CRP level on postoperative day 3.

The optimal threshold was >106 mg/L (sensitivity 85%, specificity 63%). The CRP level on day 5 also showed high value (AUC=0.77; 95% CI: 0.68–0.86) with a threshold of 60.8 mg/L (Table 2). Calibration curves for the models showed good calibration (Brier score=0.12 for the CRP-only model).

**Table 2.**
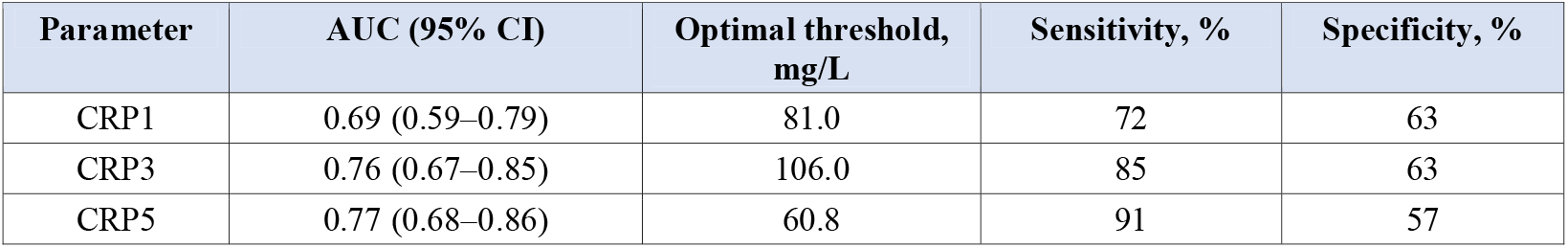
ROC analysis results for CRP levels in predicting SSI.

| Parameter | AUC (95% CI) | Optimal threshold, mg/L | Sensitivity, % | Specificity, % |
| --- | --- | --- | --- | --- |
| CRP1 | 0.69 (0.59–0.79) | 81.0 | 72 | 63 |
| CRP3 | 0.76 (0.67–0.85) | 106.0 | 85 | 63 |
| CRP5 | 0.77 (0.68–0.86) | 60.8 | 91 | 57 |

### Multivariate Analysis

Logistic regression identified the following independent predictors of SSI:

- Surgery duration (OR=1.01 per 1 minute; 95% CI: 1.01–1.02; p<0.001).
- Increase in CRP between days 3 and 5 (ΔCRP3–5) (OR=1.03 per 1 mg/L; 95% CI: 1.00–1.05; p=0.023).

Altemeier wound class III also demonstrated a clinically significant increase in risk (OR=7.10; 95% CI: 0.75–67.2; p=0.099). Full regression results are presented in Table 3.

**Table 3.**
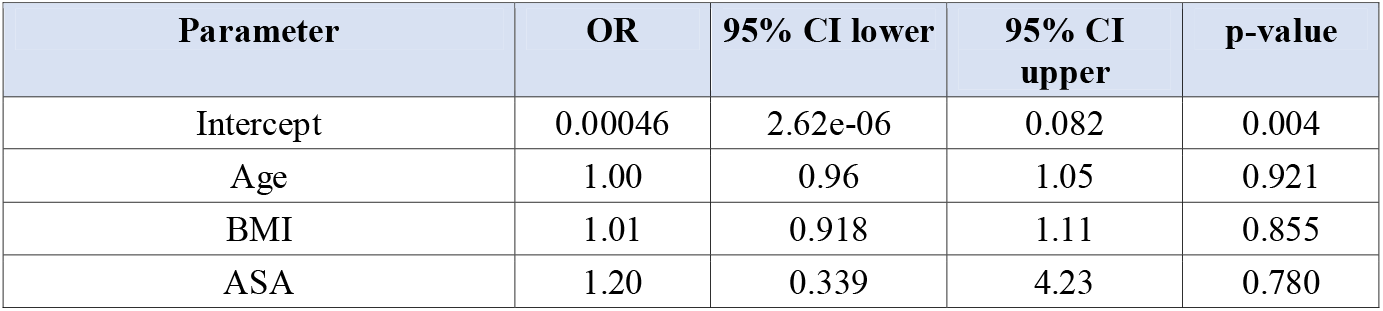
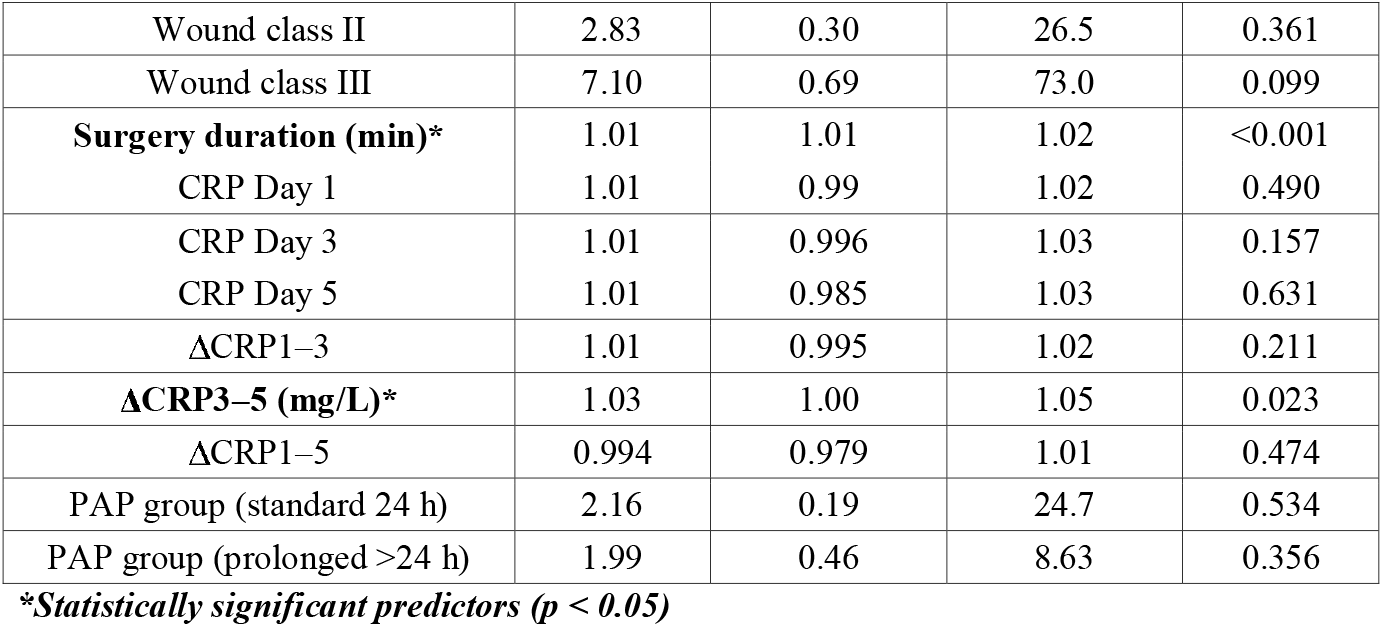
Multivariate logistic regression results for SSI predictors.

The CRP-only predictive model demonstrated high discriminatory ability (AUC=0.78; 95% CI: 0.69–0.87), which slightly increased with the addition of clinical factors in the combined model (AUC=0.79; 95% CI: 0.70–0.88) (Fig. 3).

**Figure 3.**
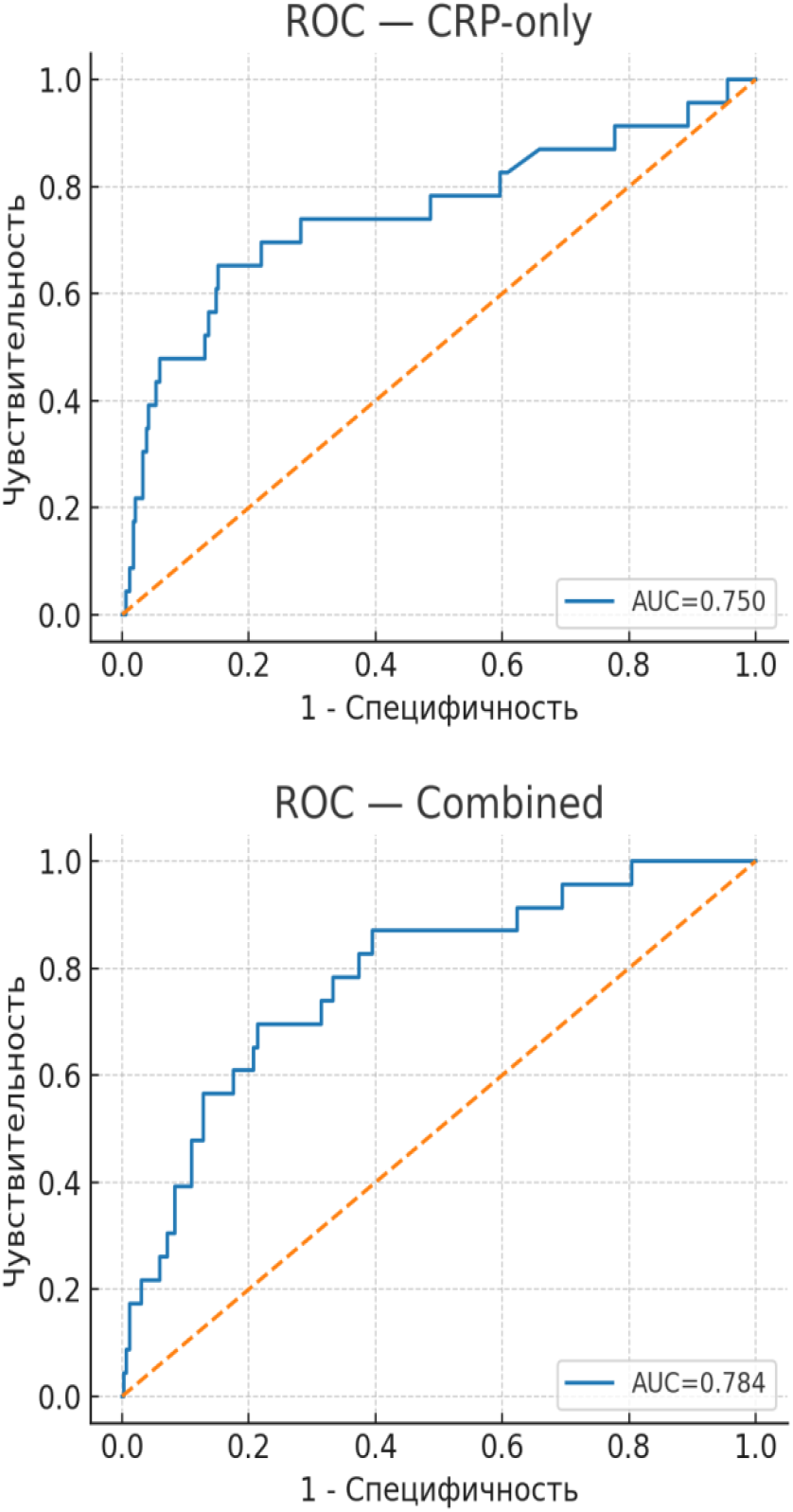
Comparison of ROC curves for prognostic models: CRP-based and combined (clinical factors + CRP). Left: CRP-only model (AUC=0.750). Right: Combined model (AUC=0.784).

### Post-hoc Power Analysis Results

A post-hoc power analysis was conducted to assess sample size adequacy and statistical power (Table 4). For comparison of CRP levels between SSI and non-SSI groups, a power of 0.82 indicates a high probability of detecting moderate differences between groups, confirming adequate sample size for detecting statistically significant effects. For multivariate logistic regression with 5 predictors, a power of 0.75 indicates an acceptable level for identifying significant predictors of SSI; however, caution is warranted in interpreting results due to a 25% risk of Type II error. This underscores the need for further validation of the model on larger samples.

**Table 4.** Post-hoc power analysis results.

| Analysis | Sample size | Effect size (Cohen's d) | $\alpha$ | Power | Conclusion |
| --- | --- | --- | --- | --- | --- |
| CRP level comparison (SSI vs. no SSI) | n=127 (21/106) | 0.5 (moderate) | 0.05 | 0.82 (82%) | High probability of detecting moderate differences |
| Multivariate regression (5 predictors) | n=127 | – | 0.05 | 0.75 (75%) | Acceptable level; caution due to 25% Type II error risk |

#### Subgroup analysis

In the emergency surgery subgroup (n=52), the CRP threshold on day 3 was >120 mg/L (AUC=0.78; sensitivity 82%, specificity 65%), while in elective cases (n=75) it was >95 mg/L (AUC=0.74; sensitivity 88%, specificity 60%). In polymorbid patients (diabetes, n=28; smoking, n=35), ΔCRP3–5 was significantly higher (median +25 mg/L vs. +10 mg/L in the overall group; p=0.045). The microbiological profile in SSI cases showed a predominance of gram-positive pathogens (Staphylococcus aureus in 45%), which correlated with a more pronounced CRP increase (r=0.42; p=0.032).

## Discussion

The results of the present study are consistent with current literature [4, 5, 8, 12–15] and confirm the high diagnostic value of monitoring CRP dynamics in the early postoperative period. The key finding is not only the significance of the absolute CRP value on day 3 (threshold >100–106 mg/L), but also the importance of its dynamics thereafter. The absence of a decline or further increase in CRP levels by day 5 (ΔCRP3–5) is a highly specific sign of a developing infectious complication and possesses independent prognostic value [4, 6, 13, 15].

The second important independent risk factor is surgery duration. An increase in surgical time by 60 minutes was associated with an approximately twofold increase in the odds ratio for SSI development, which underscores the importance of organizational measures and optimization of surgical technique for reducing infectious risks [9, 11, 14].

Post-hoc power analysis confirmed that the sample size (n=127) provides adequate power for detecting moderate differences between groups and significant predictors in the regression model. Subgroup analysis allowed accounting for potential confounders and systematic biases related to surgery type, polymorbidity, and microbiological profile, thereby enhancing the reliability of study results.

The originality of this study lies in the analysis of a local patient cohort from Krasnodar Krai, where specific features are taken into account: a predominance of mixed surgical procedures (general surgery 55%, oncology 30%, urology 15%), average procedure duration (150 min), and regional characteristics of the microbial landscape. This allows verification of patterns described by other authors (e.g., CRP thresholds of 100–150 mg/L on days 3–5 [12–15]) and identification of new trends, such as higher thresholds in emergency surgeries, possibly related to local resistance factors. Compared with alternative markers such as the CRP/albumin index (predictive value AUC=0.80 in some studies [16]) or NLR (AUC=0.75– 0.82 [6]), CRP dynamics demonstrate comparable effectiveness but are superior in accessibility and integration into routine practice, particularly in resource-limited settings.

### Clinical Implications and Recommendations

1. Determination of CRP levels on postoperative days 1, 3, and 5 in at-risk patients.
2. Assessment on day 3: CRP <100 mg/L — continue standard monitoring; CRP >100– 106 mg/L — increase clinical vigilance.
3. Assessment of dynamics by day 5: if CRP does not decline compared to day 3 (positive ΔCRP3–5), initiate an in-depth workup to rule out SSI (wound inspection, ultrasound, CT as indicated, cultures), clarify the diagnosis of bacterial infection, and only then make a decision regarding escalation of antibiotic therapy.

### Limitations

the retrospective design and the relatively small number of SSI cases (n=21) may limit the generalizability of the results. Validation of the proposed algorithms on larger prospective cohorts is required.

## Conclusions

4. Monitoring CRP dynamics within the first five days after surgery is a highly informative method for the early diagnosis of infectious complications.
5. A CRP threshold of >100 mg/L on day 3 and, more importantly, the absence of its decline or further increase by day 5 are highly sensitive markers of SSI and should serve as a trigger for an in-depth patient workup.
6. Surgery duration and CRP dynamics (ΔCRP3–5) are independent predictors of SSI development.
7. The use of CRP as the sole marker is insufficient for a definitive diagnostic conclusion; a comprehensive approach is required.
8. Application of the proposed algorithm based on CRP dynamics will enable optimization of antibiotic therapy prescribing in the early postoperative period, avoiding its unjustified use.
9. The originality of this work in the local verification of global patterns underscores the importance of regional studies for identifying trends in perioperative antibiotic prophylaxis and antibiotic therapy.

## Declarations

### Ethics statement

This study was approved by the Ethics Committee of the State Budgetary Healthcare Institution “Regional Clinical Hospital No. 2” of the Ministry of Health of Krasnodar Krai (Krasnodar, Russia). Due to the retrospective nature of the study and the use of de-identified data, the requirement for individual informed consent was waived.

### Data availability

The data supporting the findings of this study are available from the corresponding author upon reasonable request.

## Author Information

Irina N. Ochakovskaya — Department of Clinical Pharmacology and Functional Diagnostics, Kuban State Medical University; Head of the Clinical Pharmacology Department, Regional Clinical Hospital No. 2.

Vladimir V. Onopriev — Head of the Department of Clinical Pharmacology and Functional Diagnostics, Kuban State Medical University.

Narine M. Dovlatbekyan — Department of Clinical Pharmacology and Functional Diagnostics, Kuban State Medical University; Clinical Pharmacologist, Regional Clinical Hospital No. 2.

Ksenia Sh. Zhuravleva — Department of Clinical Pharmacology and Functional Diagnostics, Kuban State Medical University; Clinical Pharmacologist, Regional Clinical Hospital No. 2.

Georgiy Yu. Zamulin — Oncology Department No. 2, Regional Clinical Hospital No. 2.

Vladimir M. Durleshter — Head of the Department of Surgery No. 3, Kuban State Medical University; Deputy Chief Physician for Surgery, Regional Clinical Hospital No. 2.

